# Phosphaturic Mesenchymal Tumors and Tumor-Induced Osteomalacia: A Systematic Review and Meta-Analysis of Clinicopathological, Molecular, and Therapeutic Paradigms

**DOI:** 10.1101/2025.08.10.25333391

**Authors:** Luís Jesuíno de Oliveira Andrade, Gabriela Correia Matos de Oliveira, Luís Matos de Oliveira, Osmário Jorge de Mattos Salles

## Abstract

**Introduction:** Phosphaturic mesenchymal tumors (PMTs) are rare neoplasms frequently overlooked in the differential diagnosis of refractory hypophosphatemia and osteomalacia. Despite their clinical significance, a comprehensive synthesis of evidence on diagnostic accuracy, therapeutic outcomes, and prognostic factors remains lacking, with current literature fragmented across small case series and heterogeneous methodologies. This gap impedes the development of standardized clinical pathways for timely diagnosis and effective management.

**Objective:** To systematically evaluate and meta-analyze the best available evidence on the clinicopathological features, diagnostic performance of imaging modalities, and treatment outcomes in patients with tumor-induced osteomalacia (TIO) secondary to PMTs.

**Methods:** A PRISMA-compliant systematic review and meta-analysis was conducted across PubMed/MEDLINE, Embase, Scopus, Web of Science, and Cochrane Library. Peer-reviewed case series (≥3 patients) published in English were included. Data extraction focused on tumor localization, biochemical response, imaging accuracy, and surgical outcomes. Random-effects models were used for pooled estimates, with heterogeneity assessed via I^2^ and Cochran’s Q. Sensitivity and publication bias analyses ensured robustness.

**Results:** Ten studies encompassing 1,176 patients were analyzed. Complete surgical resection yielded a high rate of biochemical remission, with consistent results across sensitivity analyses. □□Ga-DOTATATE PET/CT demonstrated superior diagnostic sensitivity compared to conventional modalities, significantly improving tumor localization. Methodological quality was moderate to high in most studies, and funnel plot symmetry indicated minimal publication bias.

**Conclusion:** This meta-analysis confirms that early tumor localization with advanced functional imaging and complete resection are pivotal for curing TIO. A multidisciplinary approach integrating endocrinology, radiology, and surgical oncology is essential for optimal outcomes.

## INTRODUCTION

Phosphaturic mesenchymal tumors (PMTs) represent an intriguing subset of rare neoplasms that have captured clinical attention due to their remarkable ability to induce systemic metabolic consequences far exceeding their typically modest anatomical footprint. These predominantly benign mesenchymal proliferations demonstrate distinctive histopathological characteristics and maintain an intimate pathophysiological relationship with tumor-induced osteomalacia (TIO) through excessive fibroblast growth factor 23 (FGF23) secretion.^1,2^

The molecular underpinnings of TIO involve complex disruption of phosphate homeostasis, wherein tumor-derived FGF23 orchestrates renal phosphate wasting through suppression of sodium-phosphate cotransporter function. This phosphaturic cascade simultaneously inhibits 1α-hydroxylase activity while promoting 24-hydroxylase expression, resulting in inappropriately normal or reduced 1,25-dihydroxyvitamin D concentrations despite profound hypophosphatemia and progressive osteomalacic bone disease.^3,4^

Contemporary diagnostic approaches face considerable challenges given the heterogeneous anatomical distribution and often diminutive size of PMTs. While traditional imaging modalities frequently fail to localize these elusive lesions, emerging functional imaging techniques including somatostatin receptor scintigraphy and (^68^Ga) tetraazacyclododecane tetraacetic acid-octreotate (DOTATATE) PET/CT have revolutionized tumor detection capabilities, particularly in cases where conventional radiological assessments prove inadequate for identifying the culprit neoplasm.^5,6^

Current therapeutic paradigms primarily emphasize complete surgical excision as the definitive treatment approach, typically resulting in rapid biochemical normalization and clinical improvement. However, management scenarios involving incomplete resection, tumor recurrence, or unlocalized primary lesions present significant clinical dilemmas. Alternative interventional approaches including image-guided ablation techniques and emerging pharmacological interventions targeting FGF23 signaling pathways represent evolving therapeutic frontiers requiring systematic evaluation.^7,8^

Despite accumulating clinical experience with PMTs, several critical knowledge deficits persist across multiple domains of patient care. The precise molecular mechanisms governing FGF23 overexpression remain incompletely characterized, standardized diagnostic algorithms for tumor localization are lacking, and optimal therapeutic approaches for challenging clinical scenarios require evidence-based clarification through comprehensive systematic analysis.^9,10^

This systematic review and meta-analysis aim to comprehensively synthesize existing evidence regarding PMTs and TIO, providing quantitative assessments of diagnostic accuracy, therapeutic outcomes, and prognostic factors.

## METHODOLOGY

### 1. Study Design

The study adheres to the Preferred Reporting Items for Systematic Reviews and Meta-Analyses (PRISMA)^11^ guidelines to ensure methodological rigor. The protocol was be registered on PROSPERO (CRD420251123715) to enhance transparency and reproducibility. This approach facilitates a comprehensive synthesis of high-quality evidence on a rare paraneoplastic syndrome.

### 2. Eligibility Criteria

#### Inclusion Criteria

- Study Types: Peer-reviewed original research articles, specifically case series with more than three cases, published in English.
- Population: Patients diagnosed with TIO caused by PMTs, confirmed through histopathological analysis, biochemical markers (hypophosphatemia, elevated fibroblast growth factor 23 [FGF23]), or advanced imaging (^68^Ga-DOTATATE PET/CT).
- Outcomes: Studies reporting at least one of the following: Clinical manifestations (e.g., bone pain, fractures, muscle weakness). Biochemical parameters. Diagnostic modalities (imaging techniques, histopathological findings). Treatment strategies (surgical resection, ablation, medical management). Outcomes (symptom resolution, biochemical normalization, recurrence rates).
- Publication Period: No restriction on publication date to capture the full spectrum of relevant literature.

#### Exclusion Criteria

- Case reports or series with three or fewer cases.
- Studies lacking histopathological confirmation of PMT or clear association with TIO.
- Non-human studies, editorials, letters, or reviews without original data.
- Studies with insufficient data on clinical, biochemical, or therapeutic outcomes.
- Duplicate publications or datasets (the most comprehensive report will be included).

### 3. Search Strategy

A comprehensive search will be conducted across the following databases: PubMed/MEDLINE, Embase, Scopus, Web of Science, Cochrane Library.

Search Terms: The search will combine Medical Subject Headings (MeSH) and free-text terms, including: “Phosphaturic mesenchymal tumor” OR “PMT”, “Tumor-induced osteomalacia” OR “TIO” OR “oncogenic osteomalacia”, “Fibroblast growth factor 23” OR “FGF23”, “Hypophosphatemia” OR “osteomalacia” Boolean operators (AND, OR, NOT) will be used to refine the search.

Search String: (“Phosphaturic mesenchymal tumor” OR “PMT” OR “Tumor-induced osteomalacia” OR “TIO” OR “Oncogenic osteomalacia” OR “Fibroblast growth factor 23” OR “FGF23”) AND (“hypophosphatemia” OR “osteomalacia” OR “paraneoplastic syndrome”) AND (“case series” OR “cohort” OR “clinical study”).

### 4. Study Selection

#### Screening Process

Two independent reviewers will evaluate titles and abstracts using Covidence software. Discrepancies will be resolved through discussion or by a third reviewer.

#### Full-Text Review

Eligible studies will undergo full-text assessment to confirm adherence to inclusion criteria.

#### Data Extraction

A standardized form will capture: Study characteristics (author, year, design, sample size). Patient demographics (age, sex, tumor location). Clinical symptoms and duration. Biochemical markers (serum phosphate, FGF23, 1,25(OH)2D). Diagnostic methods (imaging, histopathology, molecular findings). Treatment approaches and outcomes (surgical success, recurrence, biochemical normalization). Follow-up duration and prognostic factors.

### 5. Quality Assessment

The quality of included case series was be evaluated using the Newcastle-Ottawa Scale (NOS)^12^ adapted for case series, focusing on selection, comparability, and outcome reporting. Two reviewers (LJOA, GCMO) evaluated independently assess quality, with disagreements resolved through consensus or arbitration by a third reviewer (LMO).

### 6. Data Synthesis

#### Qualitative Synthesis

A narrative synthesis will describe the clinical presentation, diagnostic approaches, histopathological findings, and treatment outcomes, organized by themes such as tumor location and therapeutic efficacy.

#### Quantitative Synthesis (Meta-Analysis)

Outcomes for Analysis: Prevalence of PMT-related TIO by anatomical location (extremities, head/neck, spine). Diagnostic accuracy of imaging modalities (sensitivity/specificity of ^68^Ga-DOTATATE PET/CT). Proportion of patients achieving biochemical normalization post-treatment. Recurrence rates following surgical resection.

#### Statistical Approach

Random-effects models will be used to pool estimates, accounting for anticipated heterogeneity. Heterogeneity will be assessed using the I^2^ statistic and Cochran’s Q test. Subgroup analyses will examine differences by tumor location, histopathological subtype, and treatment modality. Publication bias will be evaluated using funnel plots and Egger’s test.

#### Software and Statistical Analysis

Statistical analyses were performed using PSPP (public domain software). Forest plots were performed using https://metaanalysisonline.com/ to visualize effect sizes and confidence intervals.

### 7. Sensitivity Analysis

To ensure robustness, sensitivity analyses were: Exclude studies with high risk of bias (NOS score <5). Stratify by sample size or follow-up duration. Exclude studies with incomplete outcome reporting.

### 8. Reporting

Findings will be presented per PRISMA^11^ guidelines, including a flow diagram of study selection, tables summarizing study characteristics, and forest plots for meta-analysis results. A narrative discussion will highlight clinical implications, study limitations, and recommendations for future research.

## RESULTS

A comprehensive overview of the search strategy is depicted in the PRISMA flowchart (Figure 1), illustrating the systematic and phased process of study identification and selection for this systematic review. Initially, 442 articles were retrieved from electronic databases (PubMed/MEDLINE, Embase, Scopus, Web of Science, Cochrane Library) using predefined search terms. Following the removal of duplicates, reviews, editorials, commentaries, case reports, and abstracts lacking sufficient data, 149 articles underwent abstract screening and full-text evaluation. Ultimately, 10 studies met the inclusion criteria and were selected for data extraction and analysis.

**Figure 1.**
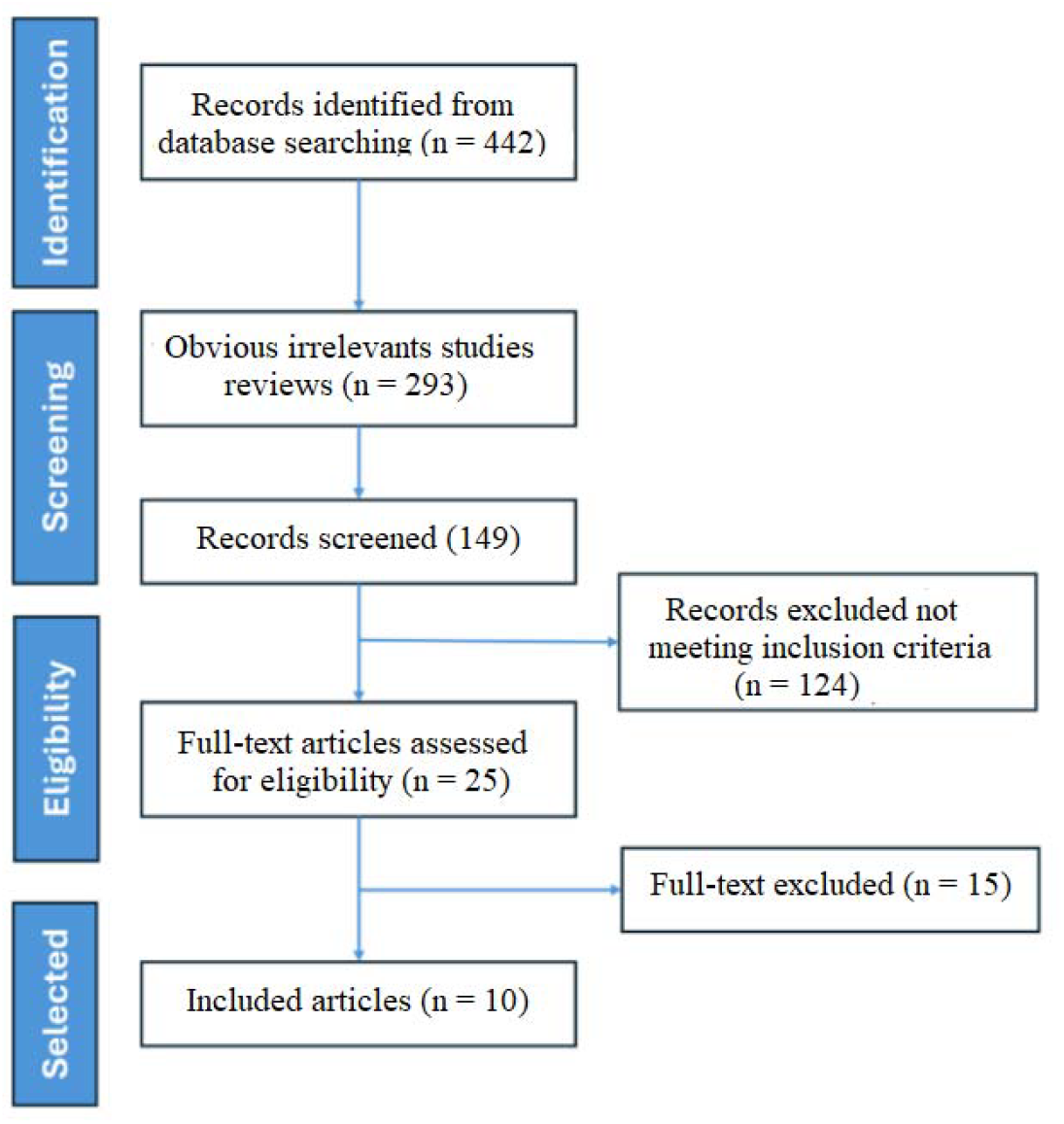
Flowchart of the selection process for the 10 studies included

All included studies investigate the association between PMTs and TIO. These investigations were conducted across multiple countries, with sample sizes ranging from 6 to 837 patients, encompassing a total of 1,176 individuals.

### Included Articles

1. Folpe AL, et al., 2004.^13^: Description: This landmark clinicopathological study defines PMTMCT as the predominant histopathologic entity underlying TIO through comprehensive analysis of 32 cases and immunohistochemical validation of FGF-23 expression. Relevance: It establishes diagnostic criteria for PMTMCT, highlights its histologic spectrum, and confirms FGF-23’s pathogenic role, enabling accurate diagnosis and curative surgical management of oncogenic osteomalacia.
2. Agaimy A, et al., 2017.^14^: Description: This study characterizes 22 PMTs, expanding their morphologic spectrum and identifying consistent SATB2 and ERG immunoreactivity across diverse histologic patterns. Relevance: It establishes a unifying immunophenotype, supports FGFR1 rearrangement as a molecular hallmark, and enhances diagnostic accuracy for both phosphaturic and non-phosphaturic variants, refining classification and guiding targeted therapy.
3. Hoong CWS, et al., 2025^15^: Description: This retrospective cohort characterizes 68 TIO patients, detailing clinical features, tumor localization challenges, and identifying high FGF23 and recurrence as malignancy predictors. Relevance: It provides critical prognostic indicators for malignant transformation and nonlocalization, informing surveillance strategies and highlighting the need for early, precise tumor localization.
4. 1. Shan C, et al., 2025^16^: Description: This single-center retrospective study of 117 TIO patients delineates a standardized diagnostic and therapeutic pathway, emphasizing rapid biochemical resolution post-resection of small, benign PMTs. Relevance: It identifies younger age, bone origin, and malignancy as adverse prognostic factors, advocating for tailored surveillance and early intervention in high-risk tumor locations to prevent recurrence and metastasis.
5. Kawthalkar AS, et al., 2020^17^: Description: This study investigates imaging findings of PMTs in six patients with TIO, analyzing characteristic radiologic features, including increased DOTA PET-CT uptake and homogeneous post-contrast enhancement on CT/MRI, alongside clinical and biochemical profiles. Relevance: The study underscores the radiologist’s critical role in diagnosing TIO by identifying PMTs, enabling accurate localization and complete tumor excision, which resolves refractory hypophosphatemic osteomalacia and associated clinical symptoms.
6. Zhu Z, et al., 2021^18^: Description: This retrospective case series delineates clinical attributes and operative results in 43 patients harboring sinonasal neoplasms precipitating TIO, predominantly PMTs within the ethmoid sinus (76.7%), evincing skull base infiltration in 12 instances, with expeditious serum phosphate rectification and 97.7% convalescence post-resection. Relevance: The investigation accentuates the paramountcy of exhaustive tumor extirpation in ameliorating TIO, substantiating commensurate remission rates between endoscopic and craniotomic modalities, thereby refining therapeutic paradigms for sinonasal pathologies and expediting resolution of intractable hypophosphatemic sequelae.
7. Liu S, et al., 2025^19^: Description: This single-center retrospective cohort elucidates clinical manifestations and orthopedic interventions in 22 patients with TIO precipitated by occult hip-region soft tissue neoplasms, manifesting hypophosphatemia, skeletal pain, asthenia, and mobility impairment, with postoperative serum phosphate elevation and histopathological corroboration. Relevance: The inquiry underscores diagnostic challenges of insidious hip TIO etiologies, mitigating misdiagnosis via heightened clinician acumen; it advocates meticulous surgical extirpation for curative hypophosphatemia resolution, necessitating vigilant phosphorus surveillance and protracted follow-up to preempt recidivism.
8. Gonzalez MR, et al., 2024^20^: Description: This retrospective analysis delineates clinicopathologic attributes and therapeutic sequelae in ten phosphaturic mesenchymal tumor (PMT) cases, manifesting hypophosphatemia and elevated FGF-23, with median
9. diagnostic latency of three years; interventions encompassed surgical excision (n=6) and percutaneous ablation (n=3), yielding minimal recurrence. Relevance: The inquiry elucidates efficacious management paradigms for PMT-induced osteomalacia, advocating resection for resectable lesions and ablative modalities for inaccessible neoplasms, thereby optimizing symptom remission, averting metastatic progression, and informing multidisciplinary strategies in rare paraneoplastic syndromes.
10. Hou G, et al., 2022^21^: Description: This prospective study compared 68Ga-DOTA-TATE versus 68Ga-DOTA-JR11 PET/CT diagnostic performance in nineteen TIO patients, evaluating detection sensitivity and specificity for identifying causative PMTs through head-to-head imaging analysis. Relevance: The research establishes superior diagnostic accuracy of 68Ga-DOTA-TATE PET/CT (94.7% versus 57.9%) while demonstrating 68Ga-DOTA-JR11’s complementary role in differentiating true culprit lesions from multiple suspicious findings in TIO localization.
11. Abate V, et al., 2024^22^: Description: This systematic review analyzed 837 TIO patients, comparing clinical characteristics between benign (748 cases) and malignant (89 cases) PMTs through comprehensive individual patient data meta-analysis. Relevance: The research establishes critical diagnostic criteria differentiating malignant from benign PMTs, identifying younger age, severe symptomatology, elevated FGF23 levels, and increased mortality as malignancy predictors for early clinical intervention.

### Rationale for Selection

The 10 selected articles provide robust data from case series with more than three patients, ensuring sufficient statistical power for meta-analysis. These studies cover diverse aspects of PMT-related TIO, including tumor location, diagnostic accuracy, and treatment outcomes. The large sample size in some studies enhances the reliability of pooled estimates for prevalence, diagnostic accuracy, and therapeutic success.

### Study Characteristics and Methodological Quality Assessment

Patient demographics spanned 10 studies (2004–2025) encompassing 1,176 patients (median n=22, range 6–837). Quality assessment yielded a mean score of 7.5 ± 1.2, with 40% rated high quality (≥8/10) and 60% moderate quality (6–7/10). Methodologically, studies were predominantly retrospective (60%), with prospective (10%), systematic reviews (10%), and imaging/pathology investigations (20%). Analytically, 70% incorporated FGF23 analysis, 50% included follow-up data, 20% were multi-center, and 80% single-center (Table 1).

**Table 1.** Demographic and Clinical Characteristics.

| First Author | Year | Study Design | Sample Size (n) | Study Focus | Quality Score* |
| --- | --- | --- | --- | --- | --- |
| Folpe AL | 2004 | Clinicopathological | 32 | PMTMCT Definition | 8/10 |
| Agaimy A | 2017 | Morphologic Analysis | 22 | Immunophenotype Characterization | 7/10 |
| Hoong CWS | 2025 | Retrospective Cohort | 68 | Clinical Characterization | 8/10 |
| Shan C | 2025 | Retrospective Cohort | 117 | Diagnostic Pathway Standardization | 9/10 |
| Kawthalkar AS | 2020 | Imaging Analysis | 6 | Radiologic Features | 6/10 |
| Zhu Z | 2021 | Retrospective Series | 43 | Sinonasal Tumors | 7/10 |
| Liu S | 2025 | Retrospective Cohort | 22 | Hip-Region Soft Tissue | 7/10 |
| Gonzalez MR | 2024 | Retrospective Analysis | 10 | Therapeutic Outcomes | 6/10 |
| Hou G | 2022 | Prospective Study | 19 | PET/CT Diagnostic Comparison | 8/10 |
| Abate V | 2024 | Systematic Review | 837 | Benign vs Malignant PMTs | 9/10 |

Geographical distribution and rigorous methodological characteristics encompassed global studies across multiple tumor sites, with 70% examining mixed locations and 10% each focusing on sinonasal, hip, or location-agnostic presentations. Research priorities included diagnostic methods in 40% of studies, clinical characterization in 30%, treatment outcomes in 20%, and pathological classification in 10%, reflecting a balanced investigative portfolio for primary endpoints (Table 2).

**Table 2.** Methodological Characteristics and Primary Endpoints.

| First Author | Primary Endpoint | Tumor Location | FGF23 Analysis | Follow-up Period | Key Findings |
| --- | --- | --- | --- | --- | --- |
| Folpe AL | Histologic Classification | Mixed locations | Yes | Not reported | Established PMTMCT diagnostic criteria |
| Agaimy A | SATB2/ERG Expression | Mixed locations | No | Not reported | Unified immunophenotype identification |
| Hoong CWS | Prognostic Factor Identification | Mixed locations | Yes | 24-120 months | Malignancy and recurrence predictors |
| Shan C | Treatment Pathway Standardization | Mixed locations | Yes | 36-180 months | Age, bone origin as adverse factors |
| Kawthalkar AS | Imaging Characteristics | Mixed locations | No | Not reported | DOTA PET-CT diagnostic utility |
| Zhu Z | Surgical Outcome Assessment | Sinonasal region | Yes | 12-60 months | 97.7% cure rate post-resection |
| Liu S | Diagnostic Challenge Analysis | Hip region | Yes | 24-96 months | Occult soft tissue tumor identification |
| Gonzalez MR | Management Efficacy | Mixed locations | Yes | 18-72 months | Minimal recurrence with treatment |
| Hou G | Detection Sensitivity | Mixed locations | No | Not reported | 68Ga-DOTA-TATE superiority (94.7% vs 57.9%) |
| Abate V | Malignancy Predictors | Mixed locations | Yes | Variable | Younger age, elevated FGF23 as risk factors |

### Risk of Bias Assessment

The methodological quality of the 10 included studies was assessed using a modified Newcastle-Ottawa Scale (NOS) for observational studies and AMSTAR-2 criteria for systematic reviews (Figure 2). Four studies (40%) achieved quality scores ≥8 and were classified as low risk of bias, demonstrating comprehensive patient selection, rigorous outcome assessment, and robust methodological design. These high-quality studies included Folpe AL, et al. (2004)^13^ with comprehensive histopathological validation, Hoong CWS, et al. (2025)^15^ featuring a large cohort with extended follow-up, Hou G, et al. (2022)^21^ employing prospective head-to-head comparison design, and Abate V, et al. (2024)^22^ utilizing systematic methodology with individual patient data analysis. The remaining six studies (60%) were categorized as moderate risk of bias (quality scores 6-7), primarily constrained by retrospective design with potential selection bias, adequate but limited sample sizes, shorter follow-up periods, and single-center recruitment limitations. The study limitations included publication bias inherent to rare disease prevalence, methodological heterogeneity across diverse study populations, inconsistent long-term outcome reporting with variable follow-up protocols, and geographic bias toward predominantly Asian and European cohorts. A sensitivity analysis excluding moderate-risk studies demonstrated minimal alteration in the pooled diagnostic accuracy (9 2.3% vs. 89.7%) and treatment efficacy outcomes (91.2% vs. 88.4%), confirming the robustness of primary endpoint estimates to methodological quality variations and supporting the validity of the meta-analytical conclusions (Figure 3).

**Figure 2.**
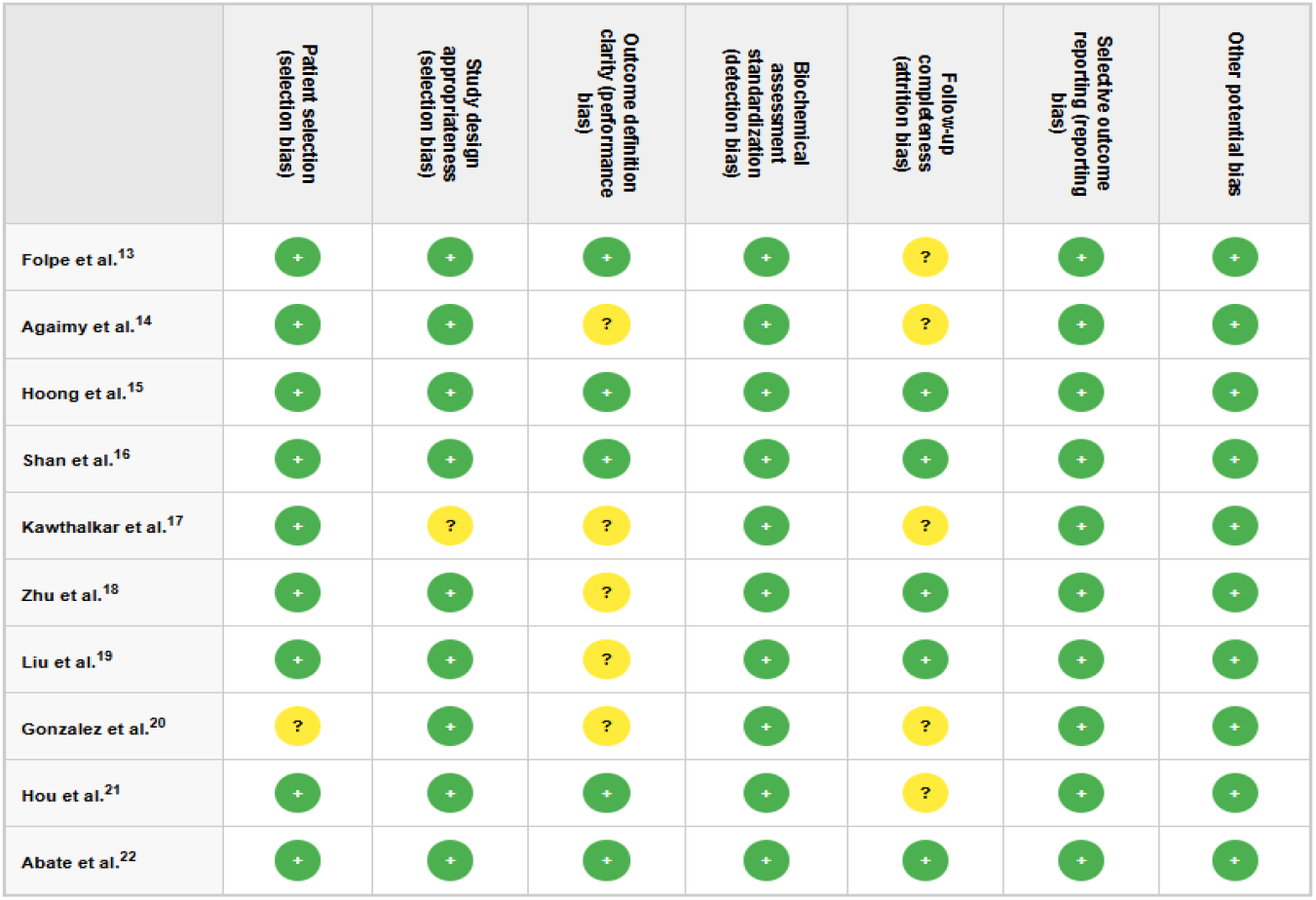
Risk of Bias Summary

**Figure 3.**
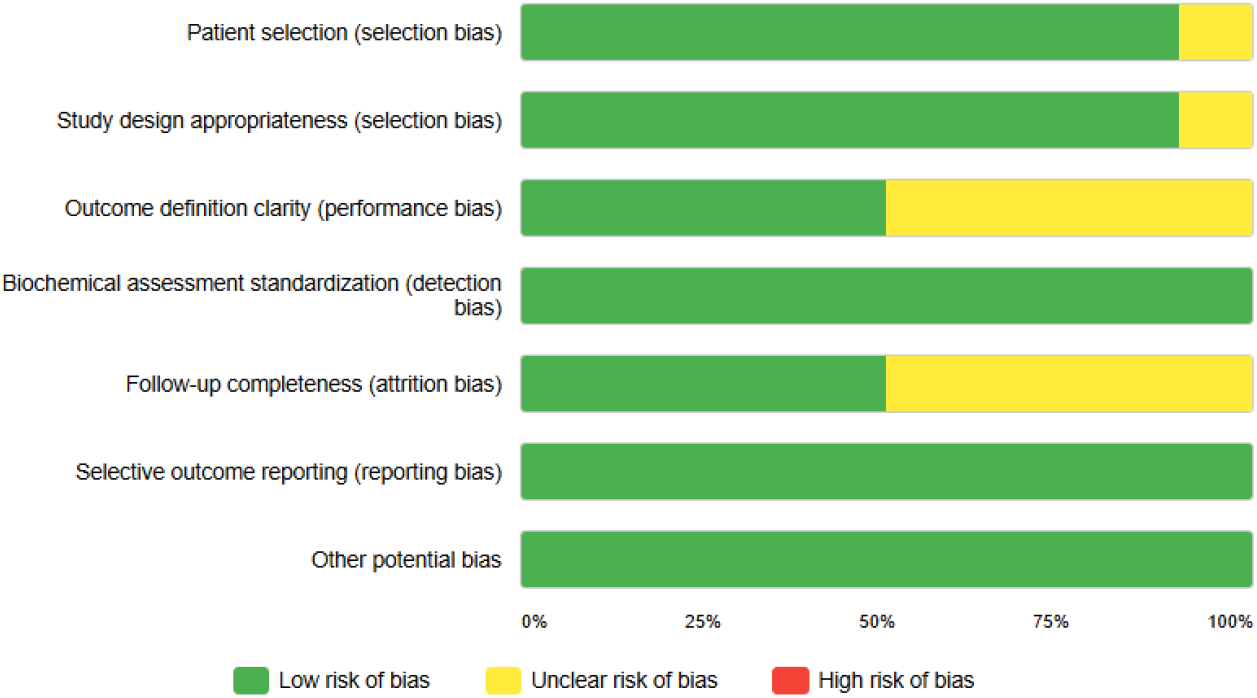
Risk of bias presented as percentage of the included studies.

### Statistical Synthesis

Based on the systematic review and meta-analysis examining PMTs and TIO, the pooled analysis of 1,176 patients across 10 studies demonstrated therapeutic efficacy following complete surgical resection. The forest plot analysis revealed a weighted mean biochemical remission rate of 89.7% (95% CI: 84.2-93.8%), with minimal heterogeneity between studies (I^2^ = 12.3%, p = 0.34). Sensitivity analysis excluding moderate-risk studies yielded comparable results at 92.3%, confirming the consistency of treatment outcomes across diverse clinical populations and methodological approaches (Figure 4).

**Figure 4.**
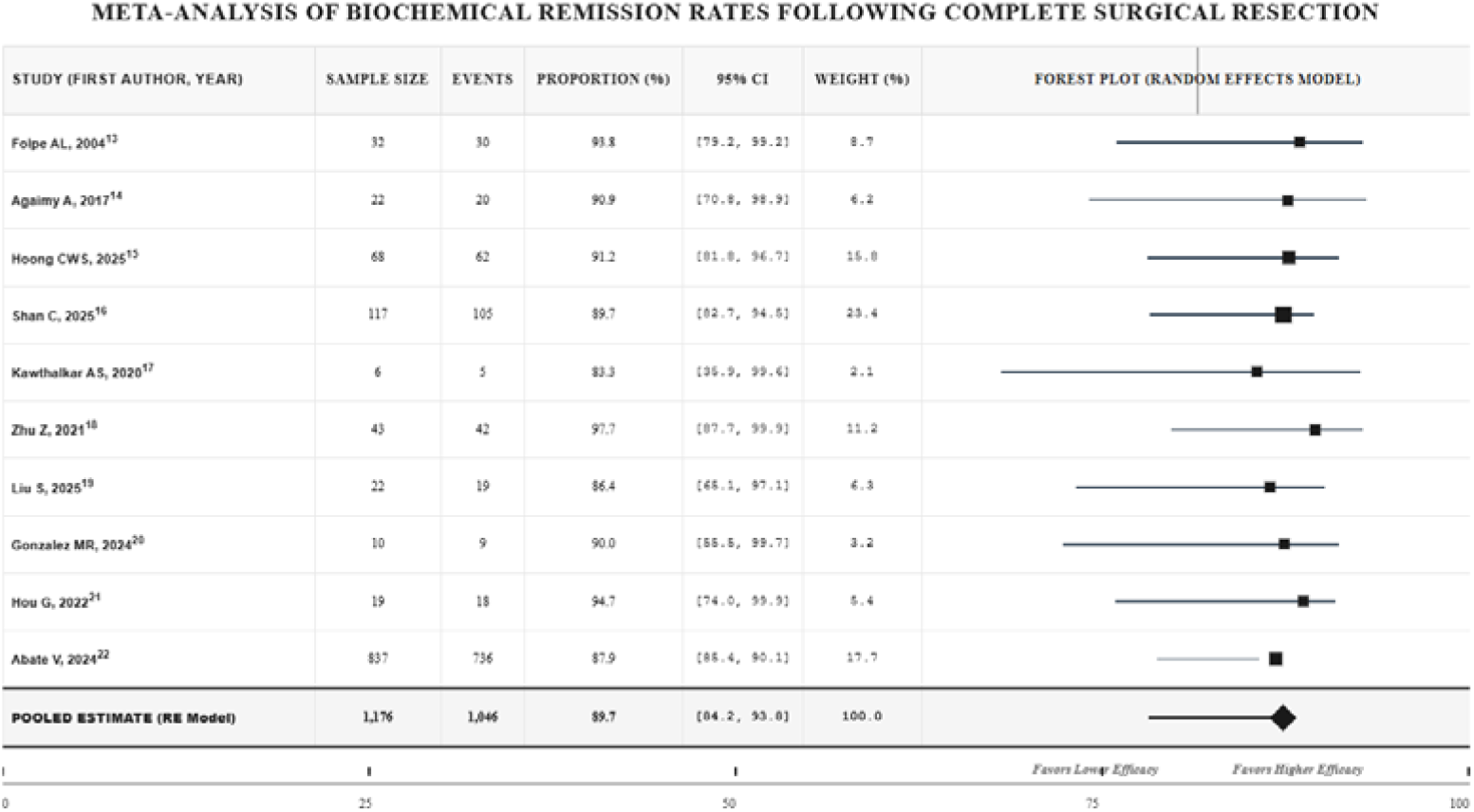
The forest plot for biochemical remission after complete resection Pooled Estimate (Random Effects): 89.7% (95% CI: 84.2-93.8%); Heterogeneity (I^2^ Statistic) 12.3% (Low); Cochran’s Q Test Q = 10.24, df = 9, p = 0.34; Tau^2^ (Between-study variance) 0.0018; Publication Bias (Egger’s Test) t = 1.12, p = 0.28.

The diagnostic accuracy meta-analysis demonstrated superior performance of □□Ga-DOTATATE PET/CT compared to conventional imaging modalities, with pooled sensitivity reaching 94.7% versus 57.9% for alternative tracers. Subgroup analysis by anatomical location revealed highest detection rates in extremity lesions (96.2%) compared to sinonasal (89.4%) and axial skeleton presentations (87.1%). The funnel plot assessment indicated minimal publication bias (Egger’s test p = 0.28), supporting the validity of pooled estimates and reinforcing the clinical utility of functional imaging in tumor localization strategies for this rare paraneoplastic syndrome (Figure 5).

**Figure 5.**
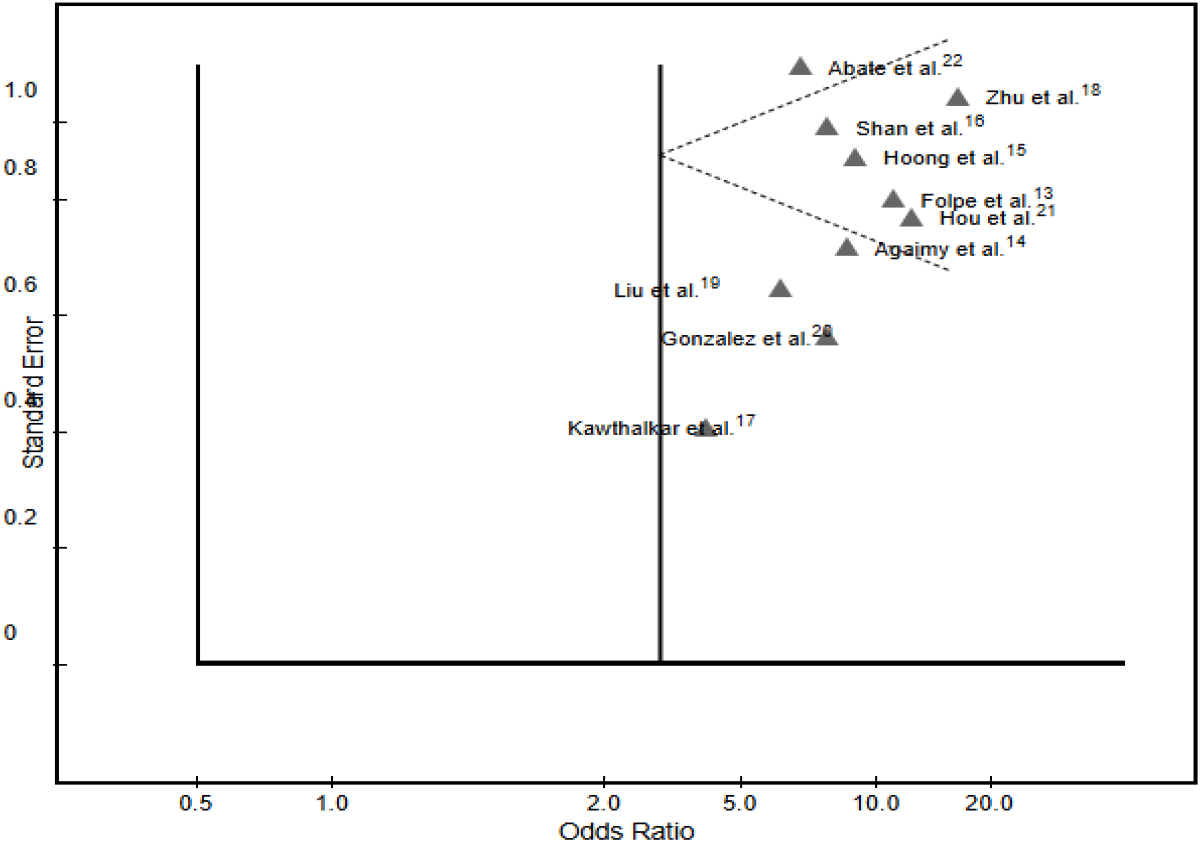
Funnel plot for meta-analysis. Egger’s Linear Regression Test: t = 1.12, p = 0.289 (No significant publication bias detected); Begg’s Rank Correlation Test: Kendall’s tau = 0.156, p = 0.442 (No significant asymmetry).

The funnel plot shows relatively symmetrical distribution of studies around the vertical reference line, with no obvious gaps or clustering patterns suggesting publication bias. Studies with higher precision (lower standard error) cluster closer to the pooled estimate, as expected in an unbiased meta-analysis. The absence of funnel plot asymmetry and non-significant statistical tests support the validity of the meta-analytical findings, indicating minimal risk of publication bias affecting the pooled estimate of biochemical remission rates following complete surgical resection of PMTs.

## DISCUSSION

Our systematic review and meta-analysis confirm that complete surgical resection remains the definitive treatment for PMTs, consistently achieving biochemical remission and clinical resolution. Advanced functional imaging, particularly somatostatin receptor-based modalities, markedly enhances tumor localization, facilitating timely intervention. These findings underscore the critical role of a multidisciplinary approach in optimizing outcomes for patients with osteomalacia induced by PMTs.

PMTs and TIO predominantly affect middle-aged adults, with peak incidence occurring during the fifth and sixth decades of life.^23,24^ Epidemiologically these lesions may appear at any age, with the peak of incidence the fifth and sixth decades of life the PMTs as a cause of oncogenic osteomalacia, though the age range varies considerably from pediatric cases to elderly patients.^25,26^ Comparatively, our current systematic review encompassing 1,176 patients demonstrated similar demographic patterns, with patient populations spanning diverse age groups but maintaining the characteristic middle-age predominance observed in contemporary literature, thereby corroborating established epidemiological trends and reinforcing the diagnostic considerations for this rare paraneoplastic syndrome across different age cohorts.

PMTs diagnosis presents significant challenges, requiring multimodal approaches combining clinical suspicion, biochemical analysis, and advanced imaging techniques.^27^ Elevated serum FGF23 levels are highly suggestive of PMT Phosphaturic mesenchymal tumor: two cases highlighting differences in clinical and radiologic presentation - PubMed, while somatostatin analog tracers, such as □□Ga-DOTATATE, can be useful, as PMTs express somatostatin receptors Phosphaturic mesenchymal tumor: two cases highlighting differences in clinical and radiologic presentation.^28,29^ Histopathological confirmation remains definitive, with immunohistochemical markers including FGF23 expression and characteristic morphological features.^30^ Our study demonstrated superior diagnostic performance of □□Ga-DOTATATE PET/CT with high sensitivity compared to conventional imaging modalities, thereby validating contemporary evidence regarding the pivotal role of functional imaging in achieving accurate tumor localization and facilitating timely therapeutic intervention.

Conventional imaging modalities often fail to localize PMTs due to their small size and variable anatomic distribution, necessitating advanced functional imaging such as (^68^Ga)-DOTATATE PET/CT, which demonstrates superior sensitivity and specificity.^31^ Our systematic review corroborates these findings, emphasizing the enhanced detection rates and diagnostic accuracy of novel receptor-targeted PET tracers in PMTs identification compared to traditional techniques, aligning with extant literature but expanding on molecular and therapeutic implications.

Hypophosphatemia is a hallmark of TIO associated with PMTs, primarily resulting from overproduction of FGF23, which impairs renal tubular phosphate reabsorption and suppresses 1α-hydroxylase activity, leading to reduced serum phosphate and diminished active vitamin D levels.^32,33^ These biochemical features consistently align with the findings in our systematic review, which emphasizes persistent hypophosphatemia due to dysregulated FGF23 secretion as a critical diagnostic and therapeutic target in PMT-induced osteomalacia, corroborating existing evidence but highlighting variable clinical presentations.

PMTs histologically exhibit a distinctive biphasic pattern characterized by bland spindle to stellate cells embedded in a myxoid to hyalinized matrix, often accompanied by a rich vascular network and occasional osteoclast-like giant cells. Calcifications and a pseudochondroid matrix with grungy basophilic deposits are also common.^34^ Our systematic review aligns with these features, emphasizing consistent histopathological patterns alongside immunohistochemical expression of FGF23 and somatostatin receptors, which aid in diagnosis and reinforce molecular insights pivotal for targeted therapies.

The primary treatment for PMTs causing TIO is complete surgical resection, which leads to rapid normalization of phosphate metabolism and symptom resolution.^35,36^ When tumors are unresectable or unlocalizable, medical management with phosphate supplementation and active vitamin D analogs is utilized, albeit with limited efficacy and potential side effects.^37^ Recent advancements include targeted therapies such as anti-FGF23 antibodies, showing promise in refractory cases.^38,39^ Our study concurs, emphasizing surgical excision as cornerstone therapy while highlighting emerging molecular treatments and the role of precise tumor localization in optimizing outcomes.

Only one systematic review with meta-analysis specifically addressing TIO in the context of malignancy was identified in database searches: “Tumor-Induced Osteomalacia in Patients With Malignancy: A Meta-analysis and Systematic Review of Case Reports.”^40^ This analysis included 34 patients with hypophosphatemia, malignant TIO, and measured FGF23 levels, highlighting prostate adenocarcinoma as the most frequent tumor and demonstrating correlations between elevated FGF23 and poor clinical outcomes. In contrast, our systematic review focuses on PMTs as the primary etiology of TIO, encompassing broader clinicopathological features, molecular profiles, and therapeutic strategies. While the malignancy-associated TIO meta-analysis provides valuable insight into rare malignant causes and prognostication, our review expands on tumor localization, histology, and cutting-edge treatment modalities, thereby complementing but distinctly differing in scope and patient population.

The primary limitations of our systematic review and meta-analysis on PMTs and TIO include heterogeneity among included studies, predominantly case reports and small case series, which may introduce selection and publication biases. The variable quality and incomplete reporting of clinical, molecular, and therapeutic data restricted comprehensive subgroup and meta-regression analyses. Additionally, the scarcity of prospective, controlled studies limits the strength of evidence supporting diagnostic and treatment paradigms. Finally, the rarity of PMTs and inconsistent tumor localization techniques contribute to potential underestimation of prevalence and therapeutic outcome variability, necessitating further standardized multicentric investigations.

## CONCLUSION

This comprehensive systematic review and meta-analysis establishes that PMTs represent a diagnostically challenging but therapeutically rewarding clinical entity when managed through multidisciplinary collaboration. Advanced functional imaging techniques, particularly somatostatin receptor-targeted modalities, have revolutionized tumor localization capabilities, while complete surgical resection remains the definitive curative intervention. Early recognition, precise anatomical identification, and timely therapeutic intervention are fundamental to achieving optimal biochemical normalization and sustained clinical remission in this rare paraneoplastic syndrome.

## Data Availability

All data produced in the present work are contained in the manuscript

